# Revamping the Day Hospital Program at North York General Hospital in response to COVID-related changes in patient demographics

**DOI:** 10.1101/2024.07.27.24310746

**Authors:** Carrol Zhou, Ishita Doval, Regina Liu

## Abstract

**Background:** The Dynamic Sustainability Framework emphasizes the need for improving programs after implementation in response to the evolving environment. This report illustrates said framework and describes significant changes made to the Psychiatric Day Hospital (DH) at North York General Hospital (NYGH) in response to pandemic-related changes in participant demographic. Patient and staff satisfaction pre- and post-program modification are compared.

**Problem:** The COVID-19 pandemic resulted in increased DH referral acuity and patient affect dysregulation. The program needed to adapt to these changes and better serve the new DH patient population.

**Methods:** DH participants and team member feedback was gathered. Five major areas of improvement were identified. Changes were systematically introduced from July 2021 to January 2022. Feedback post-implementation in 2022-2023 from patients and DH team members were gathered for comparison.

**Interventions:** Dialectical Behavioral Therapy (DBT) was adopted as the theoretical basis of the revamped Day Hospital Program. All Day Hospital staff underwent training in DBT skills, with the creation of new treatment schedules and materials. Two separate streams were created for differing patient illness severity. The program continued to run during implementation of new changes, without disruption to the existing clinical workload.

**Results:** The program transitioned from a 3-week psychoeducational and rudimentary CBT program to a dual-stream 4-week DBT-based program to address patient acuity and higher prevalence of emotional dysregulation. Quantitative and qualitative feedback from new program participants have been positive.

**Conclusions:** The Day Hospital Program at NYGH made a successful transition in response to an evolving healthcare landscape.

## Introduction

### Available Knowledge

The psychiatric Day Hospital (DH) program has been an integral part of North York General Hospital (NYGH) in Ontario, Canada for over 25 years. First introduced in Montreal in 1946, Day Hospital was a Canadian innovation that was created due to increased interest and need for psychiatric care post World War II (Cameron, 1967). It consists of intensive in-hospital psychiatric treatment and programming for multiple hours of the day, several days per week without overnight stay. Day Hospitals have since spread to other countries as they address the nuanced needs of outpatient care, support de-institutionalization, and allow decentralization of hospital-based care. The DH program serves many roles including providing access to a multidisciplinary team and a higher level of intensive care for the outpatient population. While the structure of DH programs varies, each offers flexibility and comprises a wide range of services to best address a community’s needs.

Two Cochrane reviews have demonstrated similar outcomes between patient groups admitted to hospital inpatient units and DH programs (Marshall et al., 2003; Marshall et al., 2011). In studying NYGH DH data, it was found that the cost of one day at the DH program was equivalent to 25% of the cost of one day in the inpatient unit. Additionally, DH programs give patients access to a multidisciplinary team and provide flexibility in care. The longer duration of intensive care allows for diagnostic clarification, and this treatment modality is preferred by patients over hospitalization (Guillette et al., 1978). The nature of the DH programming also reduces lengths of hospital stays and diverts potentially voluntary admissions. DH programs are valuable evidence-based treatments that support a diverse patient population across the healthcare landscape.

In the past, the NYGH DH program has been a three-week program focused on psychoeducation, behavioral activation, cultivating good sleep hygiene and basic cognitive behavioral therapy (CBT) skills. The programming ran from 9 AM to 2 PM five days a week, interspersed with long scheduled breaks, totaling 15 hours of content per week. Prior to the pandemic, the DH program demographic was diverse and came both from hospital and community referrals. Patient diagnoses ranged from depression, anxiety, bipolar/psychotic spectrum illnesses to personality disorders. The program’s strengths consisted primarily of provision of structure, routine, socialization, and access to supportive healthcare providers, with less focus on group content. Further, patients from the severe and persistent mental illness (SPMI) population would often have residual symptoms that contributed to poorer integration into the group therapies with a mixed patient population. At the time, there was no major drive nor opportunities to modernize the structure and content of the DH program.

### Problem Description

The onset of the COVID-19 pandemic in March 2020 caused all outpatient psychiatric services at North York General Hospital to shut down, with some staff redeployed to different departments.

Due to overwhelming demand, the psychiatric Day Hospital program’s referral source changed from a mixture of hospital-based and community primary care referrals to exclusively internal NYGH referrals, mainly from the inpatient psychiatric unit and emergency department. With the increase in acuity of the referrals received, the number of individuals with primary challenges of emotional dysregulation and borderline personality disorder (BPD) increased. This is unsurprising, given past research has shown that 20% of psychiatric inpatients and up to 27% of emergency visits are from patients with borderline personality disorder, while the prevalence of this diagnosis in the general population is only 1.6% (Chapman et al., 2017; Sheikh et al., 2017). More content focused groups were needed in the DH program to meet the needs of these individuals. Dialectical Behavioral Therapy (DBT), a transdiagnostic evidence-based treatment for BPD and emotional/behavioral dysregulation, would be suitable for this population. However, the allied health staff had never received any previous training in DBT and were unequipped to meet this demand. Furthermore, patients from the severe and persistent mental illness (SPMI) population continued to present with residual symptoms that contributed to poorer integration into the group therapies with mixed patient abilities. This previously recognized problem became amplified in the context of increased patient acuity. Given the changing patient population in the DH programming during the pandemic, it became apparent that the program needed to evolve to appropriately meet patient needs.

### Rationale

The most significant change in the DH patient population during the COVID-19 pandemic was the marked increase in the acuity of referrals received. It was also observed that a larger proportion of DH participants had primary challenges of emotional dysregulation or diagnosis of BPD. Feedback needed to be obtained from program participants and facilitators, as well as other Day Hospitals across the region experiencing a similar change in patient population, to devise the changes NYGH DH program needed to make to adapt to the evolving healthcare context.

### Specific Aims

The purpose of revamping the NYGH DH Program was to establish a structured program that could meet the evolving needs of the DH participants. COVID-19 caused a significant change in the patient demographic such that more content-focused programming was needed to address patient needs. The clinical presentation became highly differentiated, and the DH program needed to accommodate patients with various levels of functioning. The process of revamping the DH program, from theoretical conception to practical implementation, is explained here. Measurements for participant satisfaction before and after programmatic change are also described.

## Methods

### Context

Feedback was gathered from previous participants of the DH program via standard exit questionnaires filled by all program participants in January to September 2021, after patient demographics changed due to the pandemic, but prior to any major program change. Simultaneously, feedback was gathered from members of the treatment team via multiple in person feedback sessions in 2021. Information was also gathered from other DH programs across the province of Ontario from 2020-2021 regarding experiences with programmatic improvement and barriers to change implementation. The following areas for improvement were identified:

1. The majority of DH staff felt that more content focused groups needed to be offered in the DH program to serve the multi-diagnostic patient demographic with higher acuity of illness. From the participant surveys, only 56.1% of participants strongly agreed that they gained helpful knowledge from attending the program. (Although overall, a high percentage [89.8%] of responders still either agreed or strongly agreed that they learned helpful and relevant information.)
2. Given the prevalence of emotional and behavioral dysregulation in the recent patient population, DH facilitators felt that DBT is the best protocolized treatment to implement in the DH setting, as it is a transdiagnostic evidence-based treatment for BPD and emotional/behavioral dysregulation. Furthermore, another local DH program in Ontario, Canada had made a similar transition to DBT and found it to be readily feasible and more effective than previous programming.
3. Staff psychotherapy skills training is needed to better support patients, as 0% of allied health had any psychotherapy training in DBT prior to program change implementation, while 50% of the psychiatrists had some form of training in this modality.
4. Patients prefer a longer than current length of DH programming, with shorter breaks during the day, as only 22.4% of participants strongly agreed that the duration of programming was adequate for their needs, with 56.3% of responders either agreeing or strongly agreeing that the duration was adequate.
5. Allied health staff felt that there is a need to implement separate streams such that programming is relevant to patients with differing levels of functioning to minimize disruptions in group settings.

### Intervention(s)

The implementation process was initiated in July 2021 after the program successfully received funding from hospital leadership. An external DBT expert with experience in training staff was hired to provide DBT teaching to the treatment team. Training took place over the course of four half days, each with three hours of didactic training in DBT theory. From August 2021 to January 2022, 32 essential skills from DBT were identified that were to be adapted to the DH program. As a community of practice, two members of staff were assigned to each skill, who would learn and then teach their skill to all other staff members. The skills would then be taught to the patients in group format, as an additional session on top of current programming. Staff members also adapted original DBT handouts for the new DH program to suit the needs of the patients at NYGH. Importantly, the training and gradual introduction of the new curriculum occurred concurrently while all members of staff engaged in their normal clinical work.

Hence, the existing DH program continued to operate through the restructuring process. By January 2022, all groups and skills had been covered twice in preparation for relaunch.

A schedule was also created for two separate but parallel streams of programming in anticipation for program relaunch. Their timing was complementary such that the existing staffing could still support both streams over a full-time Monday to Friday schedule. The more content-heavy stream would comprise 19 hours of total programming per week, while the alternative stream would comprise a total of 4 hours of programming per week and contain more practical information and activities.

### Study of the Intervention(s)

Feedback was gathered from previous participants of the DH program via standard exit questionnaires filled by all participants that had completed the program in January to September of 2021 (n =189). This was the time period after the demographic of the referrals had changed, but no major changes to the DH program had been made. The survey consisted of 5 Likert scale ratings around participant satisfaction with different aspects of the program, such as the relevance of topics discussed and the duration of programming. There was also a free-text section to collect qualitative feedback about the program in general, and a section which asked the participants for the most relevant topic they learned in the DH program. A copy of the survey can be found in Appendix 1. The survey was given again to participants of the Day Hospital in 2022 (n=126) and 2023 (n=85), after the program was revamped.

### Measures & Analysis

Quantitative and qualitative feedback were inputted by researchers RL and ID. The percentage of patients scoring “agree” or “strongly agree” to prompts such as “I gained helpful knowledge and skills by attending Day Hospital” were compared pre and post implementation. Qualitatively, the groups as well as factors that patients found the most helpful to their mental health were also recorded pre- and post-program change implementation, as well as their general impression of the program.

Process measures include the percentage of staff that became trained in DBT skills by the end of the programmatic change, and balancing measures include the change in number of work hours of DH allied health staff in the program. Staff satisfaction with the new program is also reported via qualitative in-person feedback sessions.

### Ethical Considerations

There were no potential conflicts of interests identified. The participants were aware of the purpose for feedback gathering of the DH program. Given this was a quality improvement initiative, a formal ethics review process was not performed.

## Results

In April 2022, the North York General Day Hospital Program fully relaunched with 36 total beds. The programming now takes place from Monday to Friday, five days a week in four-week iterations. The programming offered is cyclical, such that any entry point resulted in the learning of the same total materials after 4 weeks of participation. The program now consists of two streams that function in a separate but simultaneous manner.

The AM stream involves patients that engage in the full Monday to Friday programming which occurs from 9 AM to 3 PM on Monday and Wednesdays, and 9 AM to 12 PM on Tuesday, Thursday, and Fridays. This stream consists largely of structured group based DBT skills teaching, with additional content on cognitive behavioral therapy (CBT), interpersonal therapy (IPT) and psychoeducational groups on health and wellness. The new program is 33.3% longer in duration than the previous DH programming.

The PM stream constitutes a smaller group of patients (up to 10 beds maximum) that are appropriate for more practical and less content-driven programming, primarily targeting the patients with severe and persistent mental illness (SPMI). This group occurs on Tuesday and Thursday afternoons from 1-3 pm, and incorporates psychoeducational groups including therapeutic recreation, pharmacy, nutrition, and peer support.

Surveys reveal that under the new program, an overwhelming majority of patients (93.7% of responders) either strongly agreed or agreed that they gained helpful knowledge and skills by attending the new DH program. 62.9% strongly agreed that they gained helpful knowledge and skills by attending the new DH program, which is an increase from 56.1% before the changes were made. 94.3% of responders agreed or strongly agreed that the information was relevant to their needs, amongst them 58.1% of responders strongly agreed to the statement. 72.0% of participants either agreed or strongly agreed that the duration of the new program was adequate to meet their needs, versus 59.5% prior to programmatic change. Overall, 92.6% of applicants agreed or strongly agreed that they were satisfied with their experience at the new DH program. 7.4% of participants felt neutral to the statement, and 0% of participants felt dissatisfied with the new DH program overall. DBT skills, specifically distress tolerance, have frequently been rated by participants as the most valuable group content in the DH program. Other than DBT skills, patients frequently mention the benefit of having structure in their day, having interpersonal interactions with others and feeling “supported and heard” by others as significant benefits of the DH programing. This was true from surveys before and after programmatic change.

100% of DH staff became trained in providing all 32 DBT skills by the end of the programmatic implementation. The total work hours of the DH allied health and psychiatrists per week remain unchanged despite significant increases in programming hours and the creation of two separate streams. Feedback from the staff during the initial programmatic change contained feelings of anxiety and uncertainty, but since the change implementation, staff feed-back has been overwhelmingly positive, as they feel more equipped to handle the challenges of higher patient acuity and emotional dysregulation and felt that the program was more relevant to patient needs. Staff also reported a higher sense of work satisfaction.

## Discussion

### Summary

Recent implementation science literature highlights the importance of understanding the evolving context of our healthcare system to continuously refine programs after initial implementation. This is also known as the dynamic sustainability framework (Chapman et al., 2017), which is exemplified by the changes implemented to the NYGH Day Hospital. Although most quality improvement literature speaks to successive small changes to optimize an existing healthcare program, at times disruptive changes are necessary to maintain a program’s relevance and efficacy especially in the face of disruptive changes to the healthcare context brought on by the pandemic.

The NYGH DH program reinvented itself to be a DBT-informed, skill focused two-stream program in the face of an increasing acuity of patients and higher prevalence of emotional dysregulation. Over 200 patients have participated in the revamped DH program since its opening. Surveys collected from patients have reported a significant positive response to the new program, as well as increased satisfaction in program length. DH allied health staff at the NYGH hospital also reported feeling more competent when dealing with the changed patient demographic, and increased pride and work satisfaction.

Had it not been for the pandemic, the impetus for such rapid change in a short period of time may not have occurred. As there were changes already occurring imposed out of necessity from the pandemic and policies of infection control, it was a ripe opportunity to create and direct the change. Additionally, as the pandemic brought about restrictions to in person care, only smaller cohorts of patients were allowed through the DH program at a time. This created opportunity and time to train existing staff in DBT and implement the new groups and changes, without overburdening the workload of allied health staff while change was being implemented.

The fact that all team members were stakeholders and active participants in program improvement created a high level of team cohesiveness and collective motivation to adapt the program to the patient demographic shift. This is especially commendable as it occurred during a pandemic that produced a significant level of burnout in many healthcare providers. The change from a single stream to two streams helped improve group dynamics and facilitate learning, especially for participants in the AM stream. The program also successfully honored patient wishes for increasing program length.

### Interpretations

Changing the programmatic content from psychoeducation and CBT to mainly DBT based caused a small increase in patient satisfaction with the relevance of information provided. Indeed, even prior to the change, the patients did not have major complaints about program content. This is likely because patients enjoyed daily structure and socialization, as well as the therapeutic environment of the DH programming. They were likely less aware of the type of material that would be most relevant to their needs even if they struggled with emotional regulation. The DH allied health, on the other hand, most acutely felt the need to pivot the program and better serve the changing patient presentations. The patients did reflect a marked increased satisfaction with the length of the program after efforts were made to prolong it. Importantly, the patients’ request for longer daily and total programming length was honored without overburdening the workload of existing DH staff.

At the time of this reporting, comparable publications studied the implementation of telemedicine during the pandemic as a response to COVID-19 disruptions to in person care (Appleton et al., 2021; Kinoshita et al., 2021; Mazziotti & Rutigliano, 2021). Our study was the first of its kind that discussed implementation of changes for the purpose of restructuring programs, while continuing to provide in-person care.

The intervention had a number of impacts on patients, staff and systems. Training and implementation did not disrupt access to DH services or care—it allowed for in-person treatment and helped provide meaningful and accessible dispositions for patients presenting to the emergency department (ED) or psychiatric inpatients who did not warrant nor benefit from admission. Patients learned new skills that were relevant and practical, particularly addressing the effects of the pandemic on mental health. For the DH clinicians, while there may have been initial resistance and anxiety to change, their investment of time and effort into learning specialized skills during and after implementation built a sense of pride and resilience. Seeing benefits to patients in the process reinforced their buy-in, and pride was amplified by being able to maintain in-person care while all other community agencies remained virtual.

Although none of the DH allied health experienced an increased workload, non-DH allied health in the hospital did have an increased workload in order to increase DH program length, such as the pharmacist and peer support workers. It may be important to solicit feedback on these individuals to see if they are satisfied with their new workload.

A trade-off of the changes is that the minority of DH program participants who are from the SPMI population are now receiving fewer total hours of programming per week compared to previous. However, these individuals are now receiving more practical programming that likely meets their learning level and are being engaged with other services at NYGH such as individual case management, which may be better catered to their needs. This is reflected by the slightly improved ratings of the program since the change implementation.

Administrative staff at the NYGH DH program have gathered demographic and diagnostic information on participants of the new program, which have not yet been analyzed. With this data, one could look closer at program utilization patterns as a potential future direction of research.

### Limitations

The program exit-surveys captured participants that had completed their entire DH programming and did not include ones that dropped out due to lack of participation. Therefore, it could have resulted in some bias in the results to be more positive than if these other participants’ opinions were collected. It may be worth considering changing the methodology to include these individuals’ feedback in the future, although that would be difficult given many patients who drop out of the program also become lost to contact.

Another limitation of the study is that data collection was limited prior to the pandemic, making it difficult to draw conclusions on how effective the original DH programming was on the original patient population.

Finally, DH is challenging to study and generalize outcomes. What can be seen as strengths, including its flexibility and catering to a transdiagnostic and heterogenous population, also creates variability that makes it difficult to study or generalize. However, measures were used in this study to compare the DH program at NYGH with itself, before and after change implementation. Similar comparisons could be made in other DH programs even if their format and content are different than the one at NYGH.

## Conclusions

The North York General Day Hospital Program made a successful transition in programmatic content to include structured DBT skills teaching. It demonstrated innovation and optimization within healthcare to serve the needs of a changing patient population. This model can be adapted for other programs looking to shift their framework of care to provide treatment for an evolving patient population.

## Data Availability

All data produced in the present study are available upon reasonable request to the authors

## Acknowledgements

The authors would like to acknowledge the following individuals:

David Koczerginski MD FRCPC

Chief of Psychiatry & Medical Director, Mental Health Program

North York General Hospital

Sandy Marangos RN, MSc

Clinical Director, Mental Health Program

North York General Hospital

who helped to make the changes implemented to the Day Hospital program possible.

## Funding Acknowledgement

Funding was received by the Mental Health Program at North York General Hospital for implementation of programmatic changes including DBT training. The authors received no financial support for the research, authorship, and/or publication of this article.

## References

Appleton, R., Williams, J., Vera San Juan, N., Needle, J.J., Schlief, M., Jordan, H., Sheridan Rains, L., Goulding, L., Badhan, M., Roxburgh, E., Barnet, t P., Spyridonidis, S., Tomaskova, M., Mo, J., Harju-Seppänen, J., Haime, Z., Casetta, C., Papamichail, A., Lloyd-Evans, B., Simpson, A., Sevdalis, N., Gaughran, F., Johnson, S. (2021) Implementation, Adoption, and Perceptions of Telemental Health During the COVID-19 Pandemic: Systematic Review. Journal of Medical Internet Research, 23(12). doi:10.2196/31746

Cameron, D. E. The origin and growth of the day hospital. (1967). Canadian Psychiatric Association Journal, 12(3), 287–291. 10.1177/070674376701200309.

Chambers, D.A., Glasgow, R.E. & Stange, K.C. (2013). The dynamic sustainability framework: addressing the paradox of sustainment amid ongoing change. Implementation Science, 117(8). 10.1186/1748-5908-8-117

Chapman, J., Jamil, R. T., & Fleisher, C. (2017). Borderline personality disorder.

Guillette, W., Crowley, B., Savitz, S. A., & Goldberg, F. D. (1978). Day hospitalization as a cost-effective alternative to inpatient care: a pilot study. Psychiatric Services, 29(8), 525–527. 10.1176/ps.29.8.525.

Kinoshita, S., Cortright, K., Crawford, A., Mizuno, Y., Yoshida, K., Hilty, D., Guinart, D., Torous, J., Correl, l C.U., Castle, D.J., Roch, a D., Yang, Y., Xiang, Y.T., Kølbæk, P., Dines, D., ElShami, M., Jain, P., Kallivayalil, R., Solmi, M., Favaro, A., Veronese, N., Seedat, S., Shin, S., Salazar de Pablo, G., Chang, C.H., Su, K.P., Karas, H., Kane, J.M., Yellowlees, P., Kishimoto, T. (2022). Changes in telepsychiatry regulations during the COVID-19 pandemic: 17 countries and regions’ approaches to an evolving healthcare landscape. Psychological Medicine, 52 (13), 2606–2613. 10.1017/S0033291720004584 [Opens in a new window

Marshall, M., Crowther, R., Sledge, W. H., Rathbone, J., & Soares-Weiser, K. (2003). Day hospital versus admission for acute psychiatric disorders. Cochrane database of systematic reviews, (12). doi: 10.1002/14651858.CD004026.

Marshall, M., Crowther, R., Sledge, W. H., Rathbone, J., & Soares-Weiser, K. (2011). Day hospital versus admission for acute psychiatric disorders. Cochrane database of systematic reviews, (12). doi: 10.1002/14651858.CD004026.pub2.

Mazziotti, R., Rutigliano, G. (2021) Tele-Mental Health for Reaching Out to Patients in a Time of Pandemic: Provider Survey and Meta-analysis of Patient Satisfaction. Journal of Medical Internet Research Mental Health, 8 (7). doi:10.2196/26187

Shaikh, U., Qamar, I., Jafry, F., Hassan, M., Shagufta, S., Odhejo, Y. & Ahmed, S. (2017). Patients with borderline personality disorder in emergency departments. Front Psychiatry, (8). 10.3389/fpsyt.2017.00136.

